# Static vs. Dynamic Cortical Thickening in Post-Stroke Recovery: A Normative Modeling Study

**DOI:** 10.64898/2026.03.12.26348297

**Authors:** Jiayouzheng Li, Yi Shan, Yinshan Wang, Chongjing Luo, Jie Xu, Jing Liu, Miao Zhang, Xinian Zuo, Jie Lu

**Author notes:** **Corresponding author:** Jie Lu **Full address:** Department of Radiology and Nuclear Medicine, Xuanwu Hospital, Capital Medical University, Beijing, China; 45 Changchun Street, Xicheng District, Beijing 100053, China. These authors contributed equally to this work (Jiayouzheng Li, Yi Shan).

## Abstract

**Background:** Subcortical stroke triggers heterogeneous cortical reorganization. We use neuroanatomical normative modeling to characterize individual differences of post-stroke cortical plasticity and resolve the ambiguity between dynamic reorganization and static traits.

**Methods:** This retrospective study included patients with acute subcortical stroke who underwent five longitudinal MRI scans and Fugl-Meyer (FM) motor assessments over 6 months. Individualized centile deviation scores for cortical thickness were computed against a normative model. Patients were stratified using spectral clustering based on baseline (<7 days) neuroanatomical profiles. Longitudinal changes in cortical thickness and their association with motor recovery were analyzed with linear mixed-effects models. We also stratified patients using raw thickness to evaluate the discriminative utility of normative model.

**Results:** A total of 65 patients (mean age, 52.7 ± 10.4 [SD]; 47 men) and 26 matched healthy controls (mean age, 52.7 ± 8.1 [SD]; 15 men) were evaluated. At baseline, the patient cohort exhibited widespread cortical thinning. Clustering revealed two distinct subgroups with similar baseline demographics and FM: Group L (*n*=50), with lower-than-normal thickness, and Group H (*n*=15), with static higher-than-normal thickness. Group L demonstrated a larger dynamic increase in contralesional cortical thickness than Group H (*β*=0.033, 95% CI 0.0029-0.063, *p*=0.03), which paralleled a faster rate of FM recovery (*β*=0.66, 95% CI 0.12-1.20, *p*=0.02). Furthermore, higher FM scores were associated with rising cortical thickness in Group L (*β*=0.21, 95% CI 0.0029-0.41, *p*=0.03), whereas FM scores tended to decrease with higher thickness in Group H (*β*=-0.10, 95% CI −0.097-0.16, *p*=0.47). Conversely, the two subgroups identified using raw thickness demonstrated no evidence of difference in the rate of recovery (*β*=0.20, 95% CI −0.63-0.23, *p*=0.37).

**Conclusions:** Active structural thickening, rather than static cortical reserve, is the important driver of motor recovery. Normative modeling distinguishes heterogeneity of stroke, providing a framework for predicting recovery potential.

## Introduction

Subcortical stroke triggers a complex cascade of neurobiological responses, leading to widespread structural reorganization beyond the primary lesion site.^1^ While cortical thickness is widely used to index this plasticity,^2,3^ the pattern of structural changes and its role in recovery remain elusive and often contradictory.^4–7^ However, a fundamental ambiguity persists in these interpretations: does a thicker cortex in a stroke patient represent a dynamic reorganization process, or merely a pre-existing, static neuroanatomical trait?^8,9^

This distinction is clinically critical yet indistinguishable in cross-sectional studies. In a single ‘snapshot’ of the brain, a patient with naturally high cortical reserve (a static trait) looks identical to a patient undergoing active compensatory thickening (a dynamic process). Therefore, to resolve the heterogeneity in stroke outcomes, it is imperative to move beyond static comparisons and dissect the longitudinal trajectories that distinguish constitutional differences from compensatory mechanisms.

Disentangling these trajectories, however, requires high-precision, individualized metrics. Traditional case-control studies are limited by their reliance on group averages, which treat inter-individual variability as noise. In a heterogeneous stroke population,^10^ this approach fails to determine whether a specific patient’s cortical thickness is ‘abnormal’ relative to their age and sex-matched peers.^11,12^ Without a standardized reference, it is impossible to accurately quantify the degree to which an individual brain deviates from its expected norm or to track how that deviation evolves.

Normative modeling provides the necessary framework to overcome these limitations.^13^ Functioning like a clinical growth chart, this approach leverages large-scale datasets of healthy individuals to establish a robust baseline of expected cortical structure. The model accounts for non-linear influences of covariates, including age and sex, which enhances statistical robustness.^14–16^ By mapping each patient against this normative model, we can convert raw thickness values into individualized deviation scores, offering a precise quantification of structural anomalies relative to the expected norm. This allows us to rigorously separate the ‘static’ background of the patient’s brain from the ‘dynamic’ pathological or restorative deviations induced by the stroke.

In this study, we combined normative modeling with a longitudinal design to investigate the heterogeneity of post-stroke motor recovery. We specifically aimed to resolve the ambiguity of cortical thickness by determining whether distinct subtypes of structural trajectories exist. By differentiating between patients who actively ‘thicken’ and those who simply ‘remain thick’, we hope to provide more precise biomarkers to predict recovery and guide personalized rehabilitation strategies.

## Methods

### Participants

The study protocol was approved by the ethics committee of Xuanwu Hospital. Written informed consent was obtained from all participants. This retrospective study enrolled patients with subcortical ischemic stroke who were scanned five times after stroke onset: within 7 days, at 14 days, 1 month, 3 months and 6 months. The healthy control group was scanned according to the same five-point schedule. Inclusion criteria for patients were: (a) stroke onset within 7 days; (b) single, unilateral subcortical infarction; (c) no prior history of other neurological or psychiatric disorders. Exclusion criteria were: (a) non-right-handedness; (b) cortical involvement of the infarction; or (c) stroke recurrence or secondary hemorrhage during follow-up.

### MRI Protocol

MRI data were acquired on a 3T MAGNETOM Tim Trio scanner (Siemens Healthcare, Erlangen, Germany) equipped with a 12-channel phased-array head coil. Structural images were acquired using a sagittal 3D-magnetization-prepared rapid acquisition gradient echo (MPRAGE) T1-weighted sequence (repetition time = 1600 ms, echo time = 2.15 ms, flip angle = 9°, voxel sizes = 1.0 × 1.0 × 1.0 mm^3^, field of view = 256 × 256).

### Neurological Assessment

To evaluate upper limb motor function, two neurologists independently administered the Fugl-Meyer (FM) assessment before and after each MRI scan. These two scores were averaged to reduce interrater differences. We utilized the 33-item upper extremity subscale, which yields a maximum score of 66. For subsequent analysis, the averaged scores were normalized to a 100-point scale. This assessment protocol did not include the lower extremity subscale.

### Preprocessing

Structural MRI data were preprocessed using the Connectome Computation System (CCS) pipeline.^17,18^ Then, cortical surface reconstruction and tissue segmentation into gray and white matter were performed using the longitudinal recon-all pipeline from FreeSurfer (v6.0.0).^19^ Finally, cortical thickness for 68 bilateral cortical regions defined by the Desikan-Killiany atlas was extracted for each participant.^20^ The mean cortical thickness for each hemisphere separately was also calculated as a weighted average of the thickness across the left and right hemispheres.

### Normative Scoring

To quantify individual deviations in thickness from normative trajectories, we computed out-of-sample centile deviations (ranging from −50 to 50) from the Lifespan Brain Chart.^13^ First, site-specific trajectories for thickness were created by adjusting the normative trajectories using data from healthy individuals only. Subsequently, the thickness centile score (ranging from 0 to 100) for each brain region was calculated for each stroke patient by comparing their data to site-specific trajectories. For instance, a centile score of 95 for the cortical thickness of the precentral gyrus indicates that the patient’s measurement exceeds that of 95% of the healthy population. To intuitively quantify deviation from the normative median, we subtracted 50 from the centile scores to yield a centile deviation score.

### Statistical Analyses

Statistical analyses were performed using R (version 4.5.2; The R Project for Statistical Computing). To investigate the change patterns in cortical thickness at baseline (<7d), a cross-sectional analysis was conducted using a linear regression model. The dependent variable was the centile deviation score for each brain region. The model incorporated five mean-centered covariates: total lesion volume and four binary variables encoding the presence or absence of a lesion in the basal ganglia, white matter, thalamus and brainstem. Significant thickness changes were identified in regions where the model’s intercept was significant (*p*<0.05, FDR corrected).

To identify patient subgroups with distinct neuroanatomical profiles, spectral clustering was applied to the matrix of baseline centile deviations.^12^ Concurrently, to evaluate the comparative discriminative utility of normative versus raw data in subgroup stratification, clustering analysis was also performed using the raw cortical thickness values.

Longitudinal changes in cortical thickness post-stroke were investigated using a linear mixed-effects multivariable regression model.^16^ The covariates were consistent with the previous model. This model incorporated a subject-specific random intercept to account for intra-subject correlations over time, with a fixed slope for the effect of time. To explore hemispheric-specific plasticity, this analysis was also performed separately on the mean cortical thickness of the ipsilesional and contralesional hemispheres.

The relationship between subgroup and motor recovery was examined using a similar linear mixed-effects model structure where FM score served as the dependent variable and the time-by-subgroup interaction was included. To mitigate potential ceiling effects, the baseline FM score was also included as a covariate.

Finally, to assess the direct association between structural changes and functional outcomes, a separate linear mixed-effects analysis was conducted within each subgroup. In this model, the FM score was the dependent variable, and the centile deviation score was included as a time-varying continuous independent variable, replacing the categorical subgroup variable.

### Data Availability

Anonymized data not published within this article will be made available by request from any qualified investigator.

## Results

### Participant Characteristics

A total of 65 consecutive patients (mean age, 52.7 ± 10.4 [SD]; 47 men) and 26 matched healthy controls (mean age, 52.7 ± 8.1 [SD]; 15 men) were included in this study. Demographic and clinical data are detailed in Table 1, and lesion details are shown in Figure 1.

**Figure 1.**
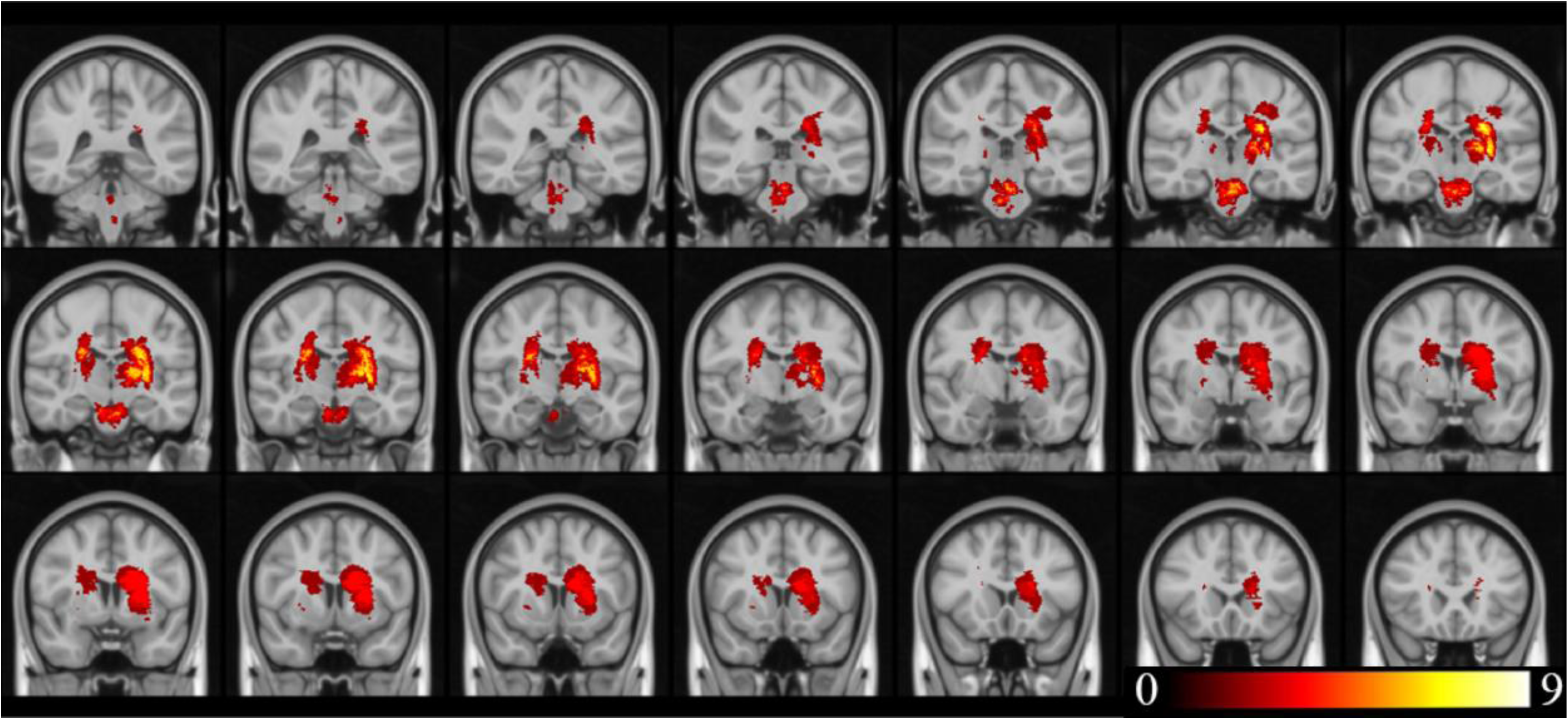
Lesion incidence map of patients with subcortical stroke. The color bar indicates the number of patients whose lesions overlap at a specific voxel.

**Table 1.**
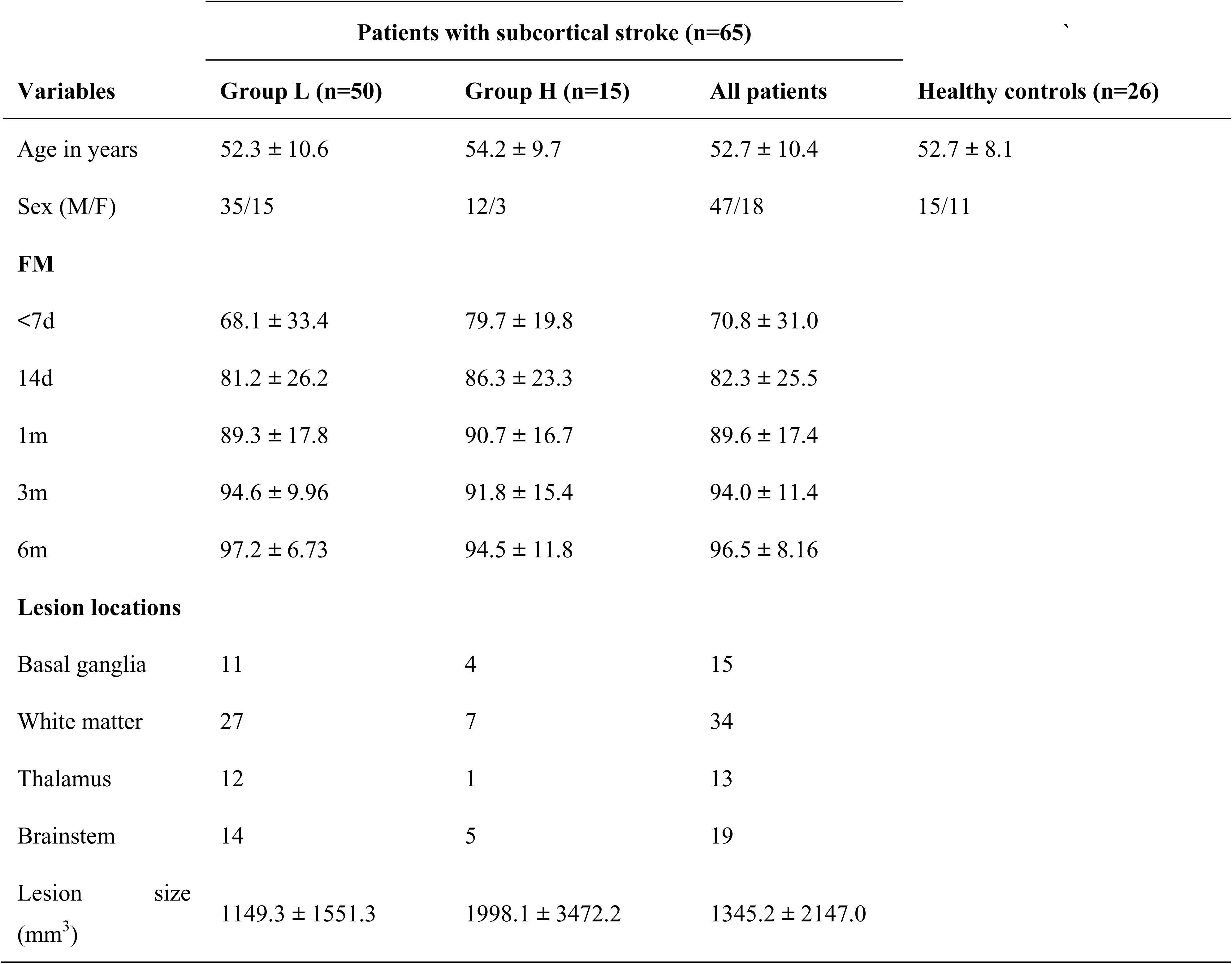
Demographic and clinical information of participants.

| Variables | Patients with subcortical stroke (n=65) |  |  | Healthy controls (n=26) |
| --- | --- | --- | --- | --- |
|  | Group L (n=50) | Group H (n=15) | All patients |  |
| Age in years | 52.3 ± 10.6 | 54.2 ± 9.7 | 52.7 ± 10.4 | 52.7 ± 8.1 |
| Sex (M/F) | 35/15 | 12/3 | 47/18 | 15/11 |
| **FM** |  |  |  |  |
| <7d | 68.1 ± 33.4 | 79.7 ± 19.8 | 70.8 ± 31.0 |  |
| 14d | 81.2 ± 26.2 | 86.3 ± 23.3 | 82.3 ± 25.5 |  |
| 1m | 89.3 ± 17.8 | 90.7 ± 16.7 | 89.6 ± 17.4 |  |
| 3m | 94.6 ± 9.96 | 91.8 ± 15.4 | 94.0 ± 11.4 |  |
| 6m | 97.2 ± 6.73 | 94.5 ± 11.8 | 96.5 ± 8.16 |  |
| **Lesion locations** |  |  |  |  |
| Basal ganglia | 11 | 4 | 15 |  |
| White matter | 27 | 7 | 34 |  |
| Thalamus | 12 | 1 | 13 |  |
| Brainstem | 14 | 5 | 19 |  |
| Lesion size (mm<sup>3</sup>) | 1149.3 ± 1551.3 | 1998.1 ± 3472.2 | 1345.2 ± 2147.0 |  |

Data are presented as the means ± SD for continuous data. Lesion locations are indicated by four binary variables encoding the presence or absence of a lesion in the basal ganglia, white matter, thalamus and brainstem, so the sum of them can exceed the number of patients. Abbreviations: FM, Fugl-Meyer assessment; M: male; F: female; Group L: low cortical thickness at baseline; Group H: high cortical thickness at baseline

### Subgroups Exhibit Two Distinct Patterns of Cortical Thickness

Application of Equation 1 to the entire patient cohort revealed widespread reductions in cortical thickness at baseline (<7d). Following the FDR correction, significant thickness reductions were observed in 44 of the 68 cortical regions analyzed (Figure 2A). Detailed information is provided in the Supplementary Table 1. The five regions exhibiting the most substantial reductions were the ipsilesional insula (*β*=-24.07, *p*<0.001, FDR corrected), the contralesional rostra middle frontal gyrus (*β*=-19.45, *p*<0.001, FDR corrected), the ipsilesional lateral orbitofrontal gyrus (*β*=-18.03, *p*<0.001, FDR corrected), the contralesional inferior temporal gyrus (*β*=-17.47, *p*<0.001, FDR corrected) and the contralesional rostral anterior cingulate (*β*=-16.71, *p*<0.001, FDR corrected).

**Figure 2.**
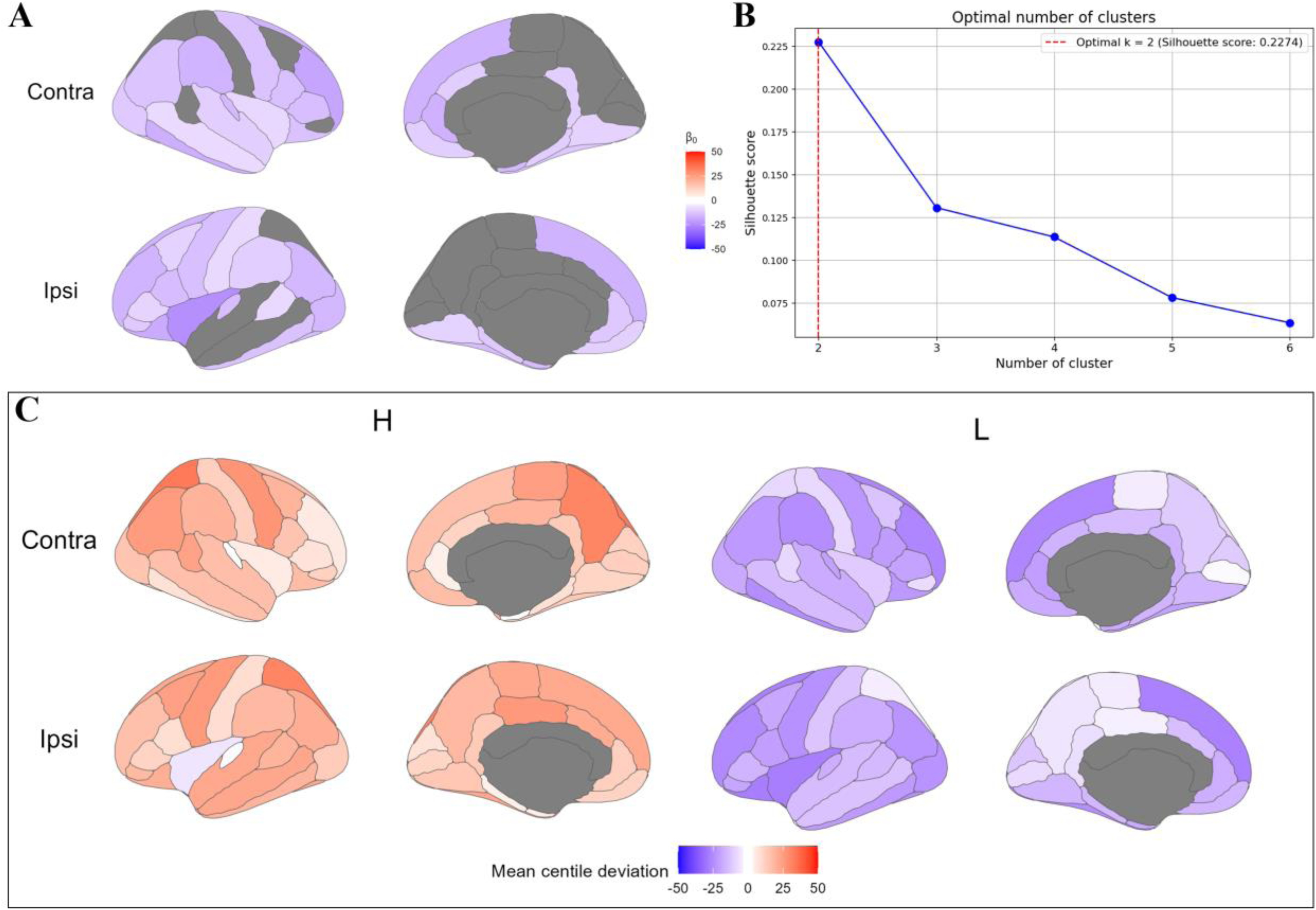
Cortical thickness centile deviation at baseline. (**A**) Brain regions showing significant (FDR corrected) non-zero cortical thickness centile deviation for all patients at baseline, as determined by the model’s intercept (𝛽_0_). The top and bottom rows depict the contralesional and ipsilesional hemispheres, respectively. (**B**) Silhouette scores across different clustering solutions (2 to 6 clusters). The optimal solution, indicated by the highest score, identified two distinct subgroups. (**C**) Mean centile deviation profiles for Group H (high cortical thickness) and Group L (low cortical thickness).

To investigate patient heterogeneity, spectral clustering was performed on the baseline cortical thickness centile deviation data. This analysis identified two distinct patient clusters, as this solution (*k*=2) yielded the highest silhouette coefficient among solutions ranging from two to six clusters (Figure 2B). The first subgroup (Group L; *n*=50) was characterized by generally lower centile deviation, whereas the second subgroup (Group H; *n*=15) displayed higher centile deviation. Due to the disparity in sample sizes, the neuroanatomical profiles of these subgroups are presented as the mean centile deviation for each brain region (Figure 2C). Importantly, no significant differences in age (*p*=0.52), sex (*p*=0.53), lesion location (*p*=0.76), lesion size (*p*=0.70), or baseline FM scores (*p*=0.35) were observed between Group L and H (Table 1).

### Longitudinal Increase in Cortical Thickness is Predominant in the Contralesional Hemisphere

A longitudinal investigation of all patients revealed a general trend of increasing cortical thickness over time. Following an FDR correction, significant increases were observed in 20 of the 68 cortical regions analyzed (Figure 3A). Detailed information is provided in the Supplementary Table 2. The five regions exhibiting the highest rates of thickening were the contralesional rostral middle frontal gyrus (*β*=0.063, *p*<0.001, FDR corrected), the contralesional lateral orbitofrontal gyrus (*β*=0.048, *p*<0.001, FDR corrected), the contralesional parsorbitalis (*β*=0.040, *p*<0.001, FDR corrected), the ipsilesional lateral orbitofrontal gyrus (*β*=0.036, *p*<0.001, FDR corrected) and the ipsilesional rostral middle frontal gyrus *(β*=0.035, *p*<0.001, FDR corrected).

**Figure 3.**
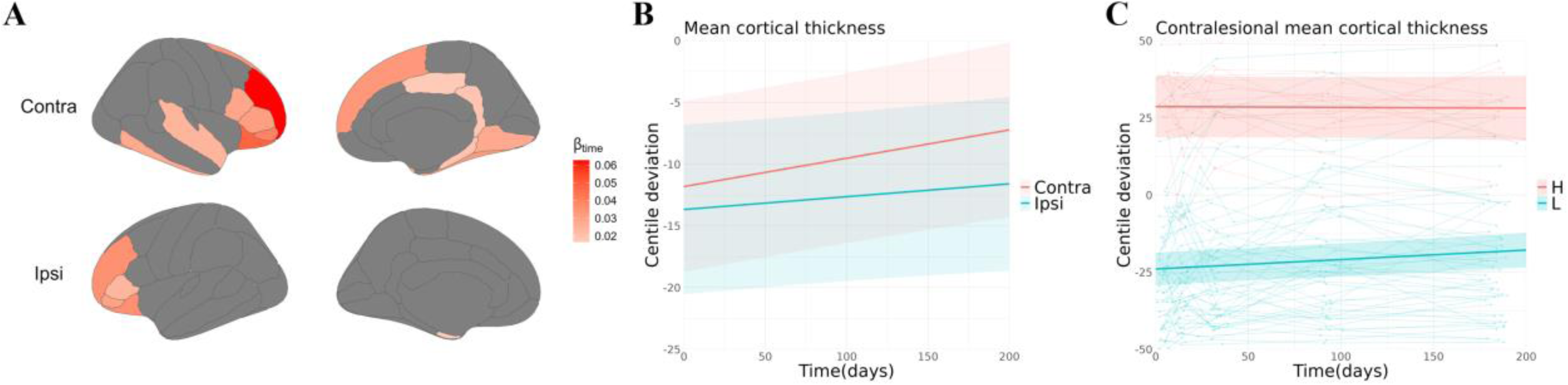
Longitudinal changes of cortical thickness centile deviation. (**A**) Brain regions showing significant (FDR corrected) changes in cortical thickness centile deviation at the whole-patient level. (**B**) Temporal changes in contralesional (red) and ipsilesional (green) mean cortical thickness at the whole-patient level. (**C**) Comparison of the rate of change in contralesional mean cortical thickness between subgroups with high (Group H, red) and low (Group L, green) baseline thickness. The semi-transparent lines in the background represent the individual centile deviation trajectories for each patient.

In terms of hemispheric-specific plasticity, the mean cortical thickness of the contralesional hemisphere showed an increase (*β*=0.023, *p*<0.001). In contrast, the ipsilesional hemisphere did not exhibit a significant change (*β*=0.010, *p*=0.12), as illustrated in Figure 3B. A cross-sectional analysis indicated no difference between contralesional and ipsilesional mean cortical thickness within 7 days post-stroke (*p*=0.44). However, at 6 months post-stroke, a difference emerged (*p*=0.005).

The two subgroups (Group L and Group H) demonstrated different patterns of cortical thickness change. While the rate of change in ipsilesional mean cortical thickness did not differ significantly between the groups, Group L exhibited a faster rate of increase in contralesional mean cortical thickness compared to Group H (*β*=0.033, 95% CI 0.0029-0.063, *p*=0.03), as illustrated in Figure 3C. Quantitatively, at baseline, the centile deviation for contralesional mean cortical thickness in Group L was 24.5 ± 17.7. At the fifth scan, this value had increased by 6.23 ± 9.87 from baseline. In contrast, Group H presented a baseline centile deviation of 80.1 ± 11.4, which subsequently decreased by 2.15 ± 7.19 at the fifth scan.

### Motor Recovery is Associated Exclusively with Dynamic Increases in Cortical Thickness

Although no significant differences in baseline FM scores were observed between Group L and H, Group L exhibited a faster rate of improvement in FM scores compared to Group H (*β*=0.66, 95% CI 0.12-1.20, *p*=0.02), as illustrated in Figure 4A. Conversely, the two subgroups identified using raw values did not demonstrate a difference in the rate of improvement (*β*=0.20, CI −0.63-0.23, *p*=0.37).

**Figure 4.**
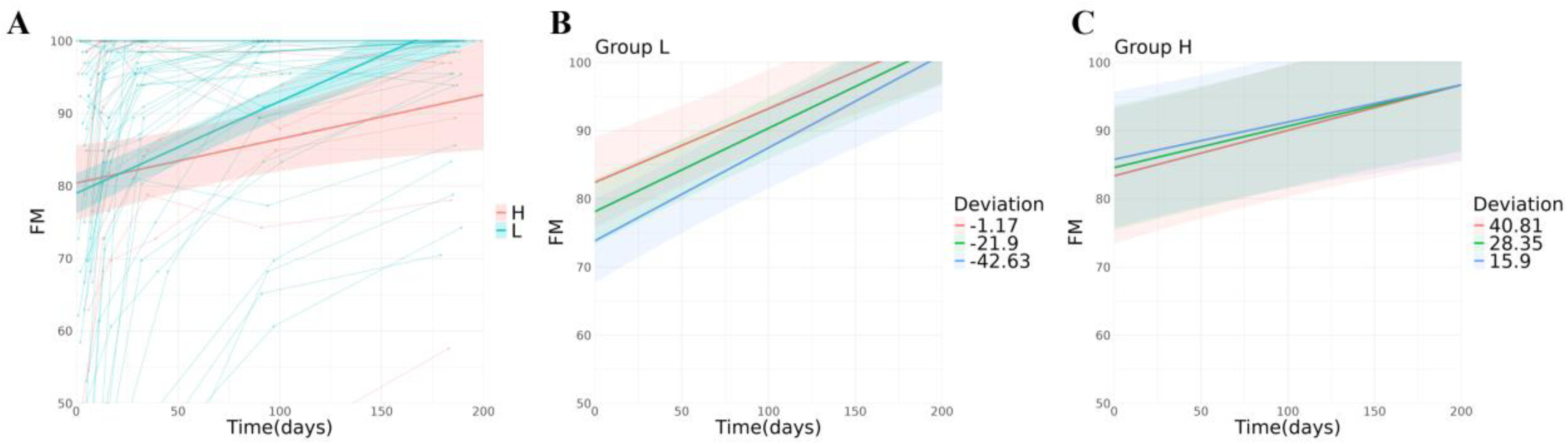
Temporal changes of FM scores. (**A**) Temporal changes in the rate of FM scores between subgroups with high (Group H, red) and low (Group L, green) baseline thickness. (**B**) The main effect of centile deviation of contralesional mean cortical thickness in Group L. The thickness shows a positive main effect on FM (𝛽_𝑑𝑒𝑣𝑖𝑎𝑡𝑖𝑜𝑛_=0.21, *p*=0.03). (**C**) The main effect of centile deviation of contralesional mean cortical thickness in Group H. The thickness shows a negative main effect on FM (𝛽_𝑑𝑒𝑣𝑖𝑎𝑡𝑖𝑜𝑛_=-0.10, *p*=0.47).

Furthermore, higher contralesional mean cortical thickness was associated with higher FM scores in Group L (*β*=0.21, 95% CI 0.0029-0.41, *p*=0.03). Still, higher thickness was associated with lower FM scores in Group H (*β*=-0.10, 95% CI −0.097-0.16, *p*=0.47), as illustrated in Figure 4B and Figure 4C. Conversely, ipsilesional mean cortical thickness did not exert a significant effect on recovery in either group. Furthermore, the interaction effects between centile deviation and time were all non-significant. Detailed information for each region is provided in the Supplementary Table 3.

## Discussion

Integrating normative modeling with longitudinal data revealed two distinct patient subgroups with divergent cortical structural profiles and motor recovery trajectories. Specifically, ‘Group L’, characterized by widespread cortical thickness below normative levels at baseline, demonstrated a significant and clinically meaningful increase in contralesional cortical thickness over time. Crucially, this dynamic thickening was directly associated with better motor recovery. In stark contrast, ‘Group H’, which maintained persistently higher-than-normal cortical thickness, exhibited slower motor recovery and a non-significant negative trend between cortical thickness and motor function.

At the whole-patient level, we observed diffuse reduction in cortical thickness followed by progressive cortical thickening over time, predominantly in the frontal and temporal lobes. This is consistent with other longitudinal studies reporting cortical thickening or volume increases in regions distant from the lesion.^4,6,21^ Our findings indicate that post-stroke cortical reorganization exhibits hemispheric specificity. Only the mean cortical thickness of the contralesional hemisphere showed a significant increase over time. This finding is consistent with a substantial body of research underscoring the critical role of the contralesional hemisphere in post-stroke recovery.^4,22–25^

To ensure that the observed disparity in recovery rates between subgroups was not an artifact of baseline functional differences (i.e., a ceiling effect where higher initial scores limit further improvement),^26^ we confirmed no significant differences in baseline FM scores, sex, age, or lesion characteristics between groups. Furthermore, we included baseline FM score as a covariate in our statistical models. Thus, we propose that the difference in cortical structural plasticity—specifically, the rate of thickness increase—drives the disparity in recovery. A dynamic increase in thickness appears beneficial, whereas a persistently high, static thickness may signify a loss of compensatory potential.

The rationale for this hypothesis is predicated on four key lines of evidence. First, during normal neurodevelopment, the nervous system undergoes synaptic pruning, where an initial overproduction of neurons, glial cells, and synapses is followed by the elimination of redundant connections, which is thought to build more efficient neural networks.^27^ Second, research has also shown that stroke risk factors, such as blood pressure and lipid levels, can themselves influence cortical thickness. For instance, the polygenic risk score for ischemic stroke has been positively correlated with cortical thickness in the occipital and parietal lobes.^28^ Moreover, cortical thickening has been observed in other pathological conditions, including psychiatric disorders,^29^ epilepsy,^30,31^ and neurodegenerative diseases like Alzheimer’s,^32,33^ where it is associated with poorer clinical outcomes. Fourth, given that the baseline difference between Group L and H was about 55.6 centile points, while the centile deviation of contralesional mean cortical thickness in Group L only increased by 6.23 centile points over 180 days, it is unlikely that the high thickness in Group H occurred solely in the acute post-stroke phase. Collectively, we hypothesize that pre-stroke factors may have contributed to the elevated cortical thickness in Group H, representing a pathological state that diminishes the potential for cortical reorganization and thereby hinders motor function recovery.

Our study has several limitations. First, its single-center design may restrict the generalizability of our findings. However, the application of normative modeling provides a basis for cross-center comparisons. Second, we lacked pre-stroke brain imaging data, which prevented us from quantifying cortical structural changes attributable to pre-existing factors. Third, the longitudinal design of the study led to the recruitment of a cohort with predominantly milder motor impairments and future research should extend this investigation to include individuals with a wider spectrum of stroke severity.

In conclusion, integrating normative modeling with longitudinal data, this study reveals that post-stroke cortical reorganization is not a uniform process but follows at least two distinct pathways with significant implications for recovery. We identified a majority subgroup of patients (Group L) who experienced faster motor recovery. A dynamic increase in contralesional thickness followed their initial cortical thinning, which contributed to improved motor function. In contrast, a smaller subgroup (Group H) with static high cortical thickness showed minimal structural change and slower functional recovery, and in this group motor function tended to be worse with higher cortical thickness. These findings suggest that the brain’s capacity for dynamic plastic reorganization, rather than a static snapshot of brain thickness, is a critical determinant of recovery. This highlights the potential of using early patient stratification and longitudinal tracking to personalize therapeutic interventions.

## Data Availability

Data generated or analyzed during the study are available from the corresponding author by request.

